# The Serological Sciences Network (SeroNet) for COVID-19: Depth and Breadth of Serology Assays and Plans for Assay Harmonization

**DOI:** 10.1101/2022.02.27.22271399

**Authors:** Amy B. Karger, James D. Brien, Jayne M. Christen, Santosh Dhakal, Troy J. Kemp, Sabra L. Klein, Ligia A. Pinto, Lakshmanane Premkumar, John D. Roback, Raquel A. Binder, Karl W. Boehme, Suresh Boppana, Carlos Cordon-Cardo, James M. Crawford, John L. Daiss, Alan P. Dupuis, Ana M. Espino, Adolfo Firpo-Betancourt, Catherine Forconi, J. Craig Forrest, Roxie C. Girardin, Douglas A. Granger, Steve W. Granger, Natalie S. Haddad, Christopher D. Heaney, Danielle T. Hunt, Joshua L. Kennedy, Christopher L. King, Florian Krammer, Kate Kruczynski, Joshua LaBaer, F. Eun-Hyung Lee, William T. Lee, Shan-Lu Liu, Gerard Lozanski, Todd Lucas, Damodara Rao Mendu, Ann M. Moormann, Vel Murugan, Nkemakonam C. Okoye, Petraleigh Pantoja, Anne F. Payne, Jin Park, Swetha Pinninti, Amelia K. Pinto, Nora Pisanic, Ji Qiu, Carlos A. Sariol, Viviana Simon, Lusheng Song, Tara L. Steffen, E. Taylor Stone, Linda M. Styer, Mehul S. Suthar, Stefani N. Thomas, Bharat Thyagarajan, Ania Wajnberg, Jennifer L. Yates, Kimia Sobhani

**Affiliations:** Department of Laboratory Medicine and Pathology, University of Minnesota, Minneapolis, Minnesota; Department of Molecular Microbiology & Immunology, Saint Louis University, Saint Louis, Missouri; Frederick National Laboratory for Cancer Research, Frederick, Maryland; W. Harry Feinstone Department of Molecular Microbiology and Immunology, The Johns Hopkins Bloomberg School of Public Health, Baltimore, Maryland; Department of Microbiology and Immunology, University of North Carolina, Chapel Hill, NC; Department of Pathology and Laboratory Medicine, Emory University School of Medicine, Atlanta, Georgia; Department of Medicine, University of Massachusetts Chan Medical School, Worcester, Massachusetts; Department of Microbiology & Immunology, University of Arkansas for Medical Sciences, Little Rock, Arkansas; Department of Pediatrics, University of Alabama at Birmingham, Birmingham, Alabama; Department of Microbiology, University of Alabama at Birmingham, Birmingham, Alabama; Department of Pathology, Icahn School of Medicine at Mount Sinai, New York, New York; Department of Pathology and Laboratory Medicine, Donald and Barbara Zucker School of Medicine at Hofstra/Northwell, Hempstead, New York; MicroB-plex, Inc., Atlanta, Georgia; Wadsworth Center, New York State Department of Health, Albany, New York; Department of Microbiology and Medical Zoology, University of Puerto Rico-Medical Sciences Campus, San Juan, Puerto Rico; Salimetrics, LLC, Carlsbad, California; Division of Pulmonary, Allergy, Critical Care and Sleep Medicine, Department of Medicine, Emory University, Atlanta, Georgia; Department of Environmental Health and Engineering, Johns Hopkins Bloomberg School of Public Health, Baltimore, Maryland; Departments of Pediatrics and Internal Medicine, University of Arkansas for Medical Sciences, Little Rock, Arkansas; Arkansas Children’s Research Institute, Little Rock, Arkansas; Department of Pathology, Case Western Reserve School of Medicine, Cleveland, Ohio; Department of Microbiology, Icahn School of Medicine at Mount Sinai, New York, New York; Virginia G Piper Center for Personalized Diagnostics, Arizona State University Biodesign Institute, Tempe, Arizona; Department of Biomedical Sciences, School of Public Health, University at Albany, Albany, New York; Center for Retrovirus Research, Department of Veterinary Biosciences, Department of Microbial Infection and Immunity, Viruses and Emerging Pathogens Program, Infectious Disease Institute, The Ohio State University, Columbus, Ohio; Department of Pathology, The Ohio State University Medical Center, Columbus, Ohio; Division of Public Health and Department of Epidemiology, College of Human Medicine, Michigan State University, East Lansing, Michigan; Unit of Comparative Medicine, University of Puerto Rico-Medical Sciences Campus, San Juan, Puerto Rico; Department of Internal Medicine, University of Puerto Rico-Medical Sciences Campus, San Juan, Puerto Rico; Center for Childhood Infections and Vaccines of Children’s Healthcare Atlanta, Department of Pediatrics, Department of Microbiology and Immunology, Emory Vaccine Center, Yerkes National Primate Research Center, Emory University School of Medicine, Atlanta, Georgia; Department of Medicine, Icahn School of Medicine at Mount Sinai, New York, New York; Department of Pathology and Laboratory Medicine, Cedars-Sinai Medical Center, Los Angeles, California

## Abstract

**Background:** In October 2020, the National Cancer Institute (NCI) Serological Sciences Network (SeroNet) was established to study the immune response to COVID-19, and “to develop, validate, improve, and implement serological testing and associated technologies.” SeroNet is comprised of 25 participating research institutions partnering with the Frederick National Laboratory for Cancer Research (FNLCR) and the SeroNet Coordinating Center. Since its inception, SeroNet has supported collaborative development and sharing of COVID-19 serological assay procedures and has set forth plans for assay harmonization.

**Methods:** To facilitate collaboration and procedure sharing, a detailed survey was sent to collate comprehensive assay details and performance metrics on COVID-19 serological assays within SeroNet. In addition, FNLCR established a protocol to calibrate SeroNet serological assays to reference standards, such as the U.S. SARS-CoV-2 serology standard reference material and First WHO International Standard (IS) for anti-SARS-CoV-2 immunoglobulin (20/136), to facilitate harmonization of assay reporting units and cross-comparison of study data.

**Results:** SeroNet institutions reported development of a total of 27 ELISA methods, 13 multiplex assays, 9 neutralization assays, and use of 12 different commercial serological methods. FNLCR developed a standardized protocol for SeroNet institutions to calibrate these diverse serological assays to reference standards.

**Conclusions:** SeroNet institutions have established a diverse array of COVID-19 serological assays to study the immune response to SARS-CoV-2 virus and vaccines. Calibration of SeroNet serological assays to harmonize results reporting will facilitate future pooled data analyses and study cross-comparisons.

## Introduction

The National Cancer Institute (NCI) Serological Sciences Network for COVID-19, or SeroNet, was launched on October 8, 2020, as a collaborative initiative to expand research on immune responses to SARS-CoV-2. SeroNet is comprised of investigators from 25 US biomedical research institutions, working in partnership with the Frederick National Laboratory for Cancer Research (FNLCR) and the SeroNet Coordinating Center, which is managed by the FNLCR.^1^ Of the 25 participating research institutions, 8 are designated as Serological Sciences Centers of Excellence (funded by U54 grants), 13 are funded with U01 grants to carry out specific research projects related to COVID-19 immunity, and 4 institutions are funded by subcontracts and are designated as Serological Sciences Network Capacity Building Centers.^1^

One of the primary goals of this partnership is “to develop, validate, improve, and implement serological testing and associated technologies.”^1^ To this end, SeroNet formed a working group, the Serology Assays, Samples, and Materials Operations Group (abbreviated as “Serology Assay Ops”), in December 2020 to allow for coordinated development and collaborative sharing of serology assay procedures, and to establish processes for harmonizing and standardizing methodologies using reference materials across institutions. Establishing harmonized and standardized SARS-CoV-2 serological assays can allow cross-comparison and pooling of research study results and facilitate clinical interpretation of results for patient care.

While there are 85 serological assays approved by the FDA for emergency use,^2^ the quick development of assays has led to the lack of harmonized cut-offs and reporting units. Furthermore, there are no consensus guidelines on reporting standards or clarity on the clinical interpretation and relevance of results. This has created a complex landscape for interpreting both research and clinical serological assay results. For example, several studies have reported on heterogeneity in serological assay performance that would have a significant impact on research study conclusions and clinical interpretations related to longitudinal serosurveillance.^3-6^ Specifically, certain assays demonstrate reduced sensitivity over time after an initial SARS-CoV-2 infection diagnosis. Muecksch et al. reported that the Abbott SARS-CoV-2 anti-Nucleocapsid IgG assay dropped from a peak sensitivity of 98% at 21 – 40 days post-PCR diagnosis, to around 70% when patients were tested ≥ 81 days post-diagnosis, whereas the Roche Elecsys SARS-CoV-2 anti-Nucleocapsid total antibody assay and Siemens SARS-CoV-2 anti-receptor-binding domain (RBD) total antibody assay both maintained high sensitivity (95 – 100%) on the same set of serial samples. Narowski et al. also found a significant decline in the longitudinal sensitivity of their lab-developed nucleocapsid assay in a study of healthcare workers.^6^ Perez-Saez et al. similarly demonstrated that the rates of sero-reversion at least 8 months after the initial infection differed greatly depending on the serological assay used.^4^ While the sero-reversion rate of the EuroImmun semiquantitative anti-S1 IgG ELISA was 26%, the rate was significantly lower for the Roche anti-Nucleocapsid total antibody assay (1.2%) and the Roche semiquantitative anti-RBD total antibody assay (0%).^4^ Additionally, numerous studies rely on neutralization assays as gold standard methods for determining the functional relevance of ligand-binding methods, but comparison studies have demonstrated variability in results for live-virus neutralization, pseudovirus neutralization, and surrogate neutralization assays (e.g., ACE2 inhibition assays),^7-9^ raising the importance of assay harmonization and standardization across laborartories.

Therefore, SeroNet aims to address these knowledge gaps in SARS-CoV-2 serological assay research by establishing collaborative initiatives to characterize, compare, and harmonize SARS-CoV-2 serological assays. This manuscript describes the depth and breadth of serological assays developed and implemented within the SeroNet consortium, and outlines a proposed process to establish assay traceability to the U.S. SARS-CoV-2 serology standard reference material and to the WHO International Standard (WHO IS 20/136) for these diverse assays, with the ultimate goal of establishing harmonized reporting standards. This will facilitate cross-comparison of results and provide clarity for their clinical interpretation, including in response to circulating SARS-CoV-2 variants.

## Methods

### Compilation of data on SeroNet serological assays

SeroNet institutions were queried by email between January and July 2021 and asked to complete a comprehensive serological assay survey to describe serological assays developed or implemented at their institution. The survey requested information on assay and sample type(s), instrument platform and reagents, data output, antibody isotype(s) detected, targeted antigens and virus strain(s), assay performance, cut-offs, use of standards and quality controls, method comparison studies, regulatory status, current use/applications for assays, and publications using each assay.

### Protocol for establishing traceability of serology assays to the U.S. SARS-CoV-2 serology standard and First WHO International Standard for anti-SARS-CoV-2 immunoglobulin

FNLCR developed a recommended protocol for SeroNet institutions to establish serology assay traceability to the U.S. SARS-CoV-2 Serology Standard. In short, for enzyme-linked immunosorbent assay platforms (ELISA), the U.S. SARS-CoV-2 standard is measured on the same 96-well plate as the daily assay standard, run as serial dilutions in triplicate and quadruplicate respectively (**Figure 1**). Standard curves are constructed for both the U.S. SARS-CoV-2 Serology standard and daily assay standard. A test of parallelism and linearity between the two dose-response curves is then performed to ensure that immunoaffinity differences or matrix effects do not prevent accurate calibration with the U.S. SARS-CoV-2 Serology Standard. Units based on the U.S. SARS-CoV-2 serology standard can then be assigned to the assay daily standard, to harmonize assays and units for results reporting. For non-plate-based assay platforms, similar dilution-based standard curves are constructed.

**Figure 1:**
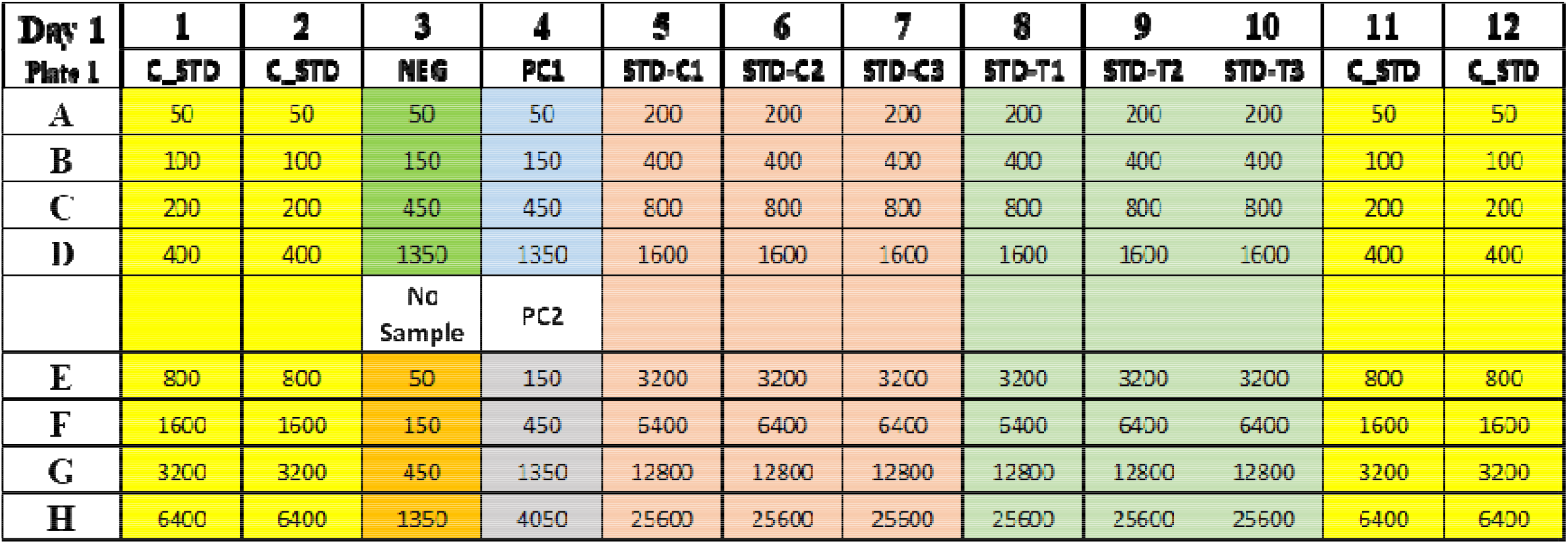
Example plate map for assay calibration set-up Numbers indicate suggested serial dilutions. Serial dilutions of primary and secondary calibrators (reference materials) are plated in triplicate, and the daily internal assay standard is plated in quadruplicate. C_STD: Daily internal assay standard STD-C1, C2, and C3: Primary calibrator (primary reference material or standard) STD-T1, T2, and T3: Secondary calibrator (secondary reference material or standard) NEG: Negative control sample PC1: Positive control sample 1 PC2: Positive control sample 2

Traceability of the FNLCR standard to the First WHO International Standard (IS) for anti-SARS-CoV-2 immunoglobulin (20/136) was established, to allow SeroNet assays to convert U.S. Serology Standard units to WHO IS units. The WHO IS 20/136 is a freeze-dried equivalent of 0.25 mL of pooled plasma from 11 individuals with a history of SARS-CoV-2 infection. Once reconstituted the WHO standard has an arbitrary unitage of 1000 binding antibody units (BAU)/mL. Eight serial dilutions of the U.S. SARS-CoV-2 serology standard and WHO IS 20/136 were run in triplicate. Parallel line analysis, which included tests for parallelism and linearity, was utilized to assign WHO IS 20/136 standard units to the U.S. SARS-CoV-2 serology standard; this will allow SeroNet institutions to convert U.S. SARS-CoV-2 serology standard units to WHO standard units for serological methods.

## Results

### SeroNet Serology Assay data

Of the 25 institutions involved with SeroNet, 23 institutions reported performing between one to seven serology assays, and provided descriptive and performance data. Serology assay data were also obtained from the Frederick National Laboratory for Cancer Research (FNLCR) and National Institute of Standards and Technology (NIST), both of which collaborate with SeroNet. Collectively, Seronet institutions reported development of 27 in-house ELISA methods (**Table 1**).^6,10-26^ The majority of ELISA methods were developed for testing of serum and/or plasma, with additional methods available for testing dried blood spots (DBS), saliva/oral fluid, and breast milk. Two methods have been granted FDA EUA approval, 3 methods are pending FDA EUA, 4 methods are validated for high-complexity testing in a CLIA-certified laboratory, and 18 methods are for research-use only (RUO). Diagnostic sensitivity and specificity for in-house ELISA methods ranged from 67.4 – 100 % and 90 – 100%, respectively.

**Table 1:** Laboratory-developed singleplex ELISA assays

| Sample Type | Antigen | Isotype | Assay Sensitivity & Specificity | Center/Institution | References | Regulatory Status |
| --- | --- | --- | --- | --- | --- | --- |
| Serum, Plasma, Dried Plasma samples | RBD | IgG (IgA/IgM being eval) | Day 0-7 after infection: Sensitivity 73.01%; Day 8-14 after infection: Sensitivity 100%; Day ≥ 15 after infection: Sensitivity 100%; Specificity (n=388 samples collected prior to COVID-19 pandemic): 97.68% | Emory University | PMID: 32835303 | FDA EUA granted |
| Serum, Plasma | RBD and Spike | IgG, IgM, IgA | Sensitivity 95%, Specificity 100% (n=38 positive, n=74 negative sera tested) | Mount Sinai | PMID: 32302069, PMID: 32511441, PMID: 33142304 | FDA EUA granted |
| Serum, Plasma, Saliva | RBD | Total Ig, with IgG, IgM, IgA titers | Overall sensitivity 82.5%, overall specificity 100% (n=300); At > 14 days from symptom onset, sensitivity 100%, specificity 100% (n=261); | University of Minnesota | PMID: 32791053, PMID: 33539808 | Assays validated in a high-complexity testing CLIA laboratory |
| Serum, Plasma | RBD | IgG, IgM | Sensitivity: 91% for RBD IgG 15-21 days post onset of symptoms, 100% >21 days post-onset of symptoms; 90% for RBD IgM 15-21 days post onset of symptoms, 100% >21 days post-onset of symptoms. Specificity: 99.75% for RBD IgG, 100% for RBD IgM | Stanford University | PMID: 33288645 | Assays validated in a high-complexity testing CLIA laboratory |
| Serum, Plasma | RBD-ACE2 | Total IgG that blocks RBD-ACE2 binding | N/A, used as a follow-up assay in seropositive specimens | Stanford University | PMID: 33288645 | Assay validated in a high-complexity testing CLIA laboratory |
| Serum, Plasma | RBD | IgG, IgM + IgG | Sensitivity 98% (n=181), Specificity 98.9% (n=181). | University of Puerto Rico | PMID: 34696403, https://www.biorxiv.org/content/10.1101/2020.06.11.146332v2 | Assay validated in a high-complexity testing CLIA laboratory |
| Serum, Plasma | Spike | IgG | Sensitivity 98.3% (n=60), Specificity 99.3% (n=150) | Frederick National Laboratory | NR | RUO |
| Serum, Plasma | Spike | IgM | Sensitivity 93.8% (n=30), Specificity 97.6% (n=80) | Frederick National Laboratory | NR | RUO |
| Serum, Plasma | Nucleocapsid | IgG | Sensitivity 97% (n=34), Specificity 100% (n=99) | Frederick National Laboratory | NR | RUO |
| Serum, Plasma | Nucleocapsid | IgM | NR | Frederick National Laboratory | NR | RUO |
| Serum, Plasma, Saliva | RBD | Total Ig | Sensitivity 95% (n=259; 9 or more days after symptom onset), Specificity 96% (n=535) | University of North Carolina | PMID: 32527802, 35090596 | FDA EUA pending |
| Serum, Plasma, Saliva | Spike NTD | Total Ig | Sensitivity = 92% (n=259; 9 or more days after symptom onset), Specificity = 94% (n=535) | University of North Carolina | PMID: 35090596 | FDA EUA pending |
| Serum | Spike, RBD | IgG | NR | CVVR/BIDMC/Harvard | PMID: 34107529 | RUO |
| Serum, Plasma, Breast milk | RBD | IgG, IgA, IgM | NR | CVVR/BIDMC/Harvard | PMID: 33983379, PMID: 33893169 | RUO |
| Serum, Plasma | Spike | IgG | Sensitivity 100%, Specificity 98.8% | Tulane University | NR | RUO |
| Serum, Plasma | RBD | IgG | NR | Tulane University | NR | RUO |
| Serum, Plasma | Nucleocapsid | IgG | NR | Tulane University | NR | RUO |
| Plasma, Serum | Spike, RBD | IgM, IgG, IgA | Spike: IgG (Sensitivity 96.6%, Specificity 96.7%); IgA (Sensitivity 99.3%, Specificity 90%); IgM (Sensitivity 97.9%, Specificity 100%). RBD: IgG (Sensitivity 97.3%, Specificity 100%); IgA (Sensitivity 99.3%, Specificity 96.7%); IgM (Sensitivity 97.9%, Specificity 96.7%). IgG data based on n=126 convalescent plasma donors, n=30 pre-pandemic samples; IgM/IgA data based on n=20 hospitalized, n=30 pre-pandemic samples. | Johns Hopkins University | PMID: 32764200 | RUO |
| Serum, Plasma | Spike (ECD), RBD | IgG | NR | University of Texas-Austin | PMID: 32910806 | RUO |
| Serum, Plasma | RBD | IgG | Sensitivity 100% (n=155), Specificity 96.5% (n=133) | Arizona State University | NR | RUO |
| Serum, DBS | RBD | IgG, IgM | Sensitivity 97% (n=39), Specificity 100% (n=37) | University of Arkansas for Medical Sciences | PMID: 34478478, https://www.medrxiv.org/content/10.1101/2021.08.04.21261592v3 | RUO |
| Serum, DBS | RBD, Spike, Nucleocapsid | IgG, IgM | Sensitivity 97% (n=39), Specificity 100% (n=37) | University of Arkansas for Medical Sciences | PMID: 34478478, https://www.medrxiv.org/content/10.1101/2021.08.04.21261592v3 | RUO |
| Serum, Plasma, Breast milk | RBD, Spike, Nucleocapsid | IgG, IgM, IgA | 97% Sensitivity (n=114), Specificity 99% | University of Alabama-Birmingham | NR | RUO |
| Serum, Plasma | RBD, Nucleocapsid, Spike Trimer | IgG, IgA | RBD: Sensitivity (70.9% for IgG, 74.4% for IgA) and Specificity (100% for both IgG and IgA); Nucleocapsid: Sensitivity (81.4% for IgG, 77.9% for IgA) and Specificity (98.5% for IgG, 100% for IgA); Spike Trimer: Sensitivity (67.4% for both IgG and IgA) and Specificity (98.5% for IgG, 100% for IgA). Data based on PCR confirmed COVID-19 hospitalized patients (n=86) and negative pre-pandemic samples (n=65). | University of Massachusetts Chan Medical School | PMID: 32780998 | RUO |
| Serum, Plasma | Nucleocapsid | IgG | Sensitivity 100% (n=44), Specificity 99.5% (n=202) | The Ohio State University | PMID: 33035201 | FDA EUA pending |
| Serum | Nucleocapsid | IgG | NR | The Ohio State University | NR | RUO |
| Oral fluid | Nucleocapsid | IgG | Sensitivity 92% (n=24), Specificity 98% (n=85) | Salimetrics | NR | RUO |

ACE2: Angiotensin converting enzyme-2; BIDMC: Beth Israel Deaconess Medical Center; CLIA: Clinical Laboratory Improvement Amendments; CVVR: Center for Virology and Vaccine Research; DBS: Dried blood spots; ECD: Extracellular domain; EUA: Emergency Use Authorization; FDA: Food and Drug Administration; NR: Not reported; NTD: N-terminal domain; PMID: PubMed Identifier; RBD: receptor binding domain; RUO: research use only

Eight institutions reported development or use of multiplex or protein arrays for antibody detection (**Table 2**).^27-37^ Sample types include serum, plasma, DBS, saliva, and bronchoalveolar lavage (BAL) fluid. Diagnostic sensitivity and specificity for multiplex and protein array methods range from 85 – 98.8 % and 95.2 – 100 %, respectively. Neutralization assays were developed by 9 institutions, with sample types including serum, plasma, BAL fluid, nasal wash, DBS, and breast milk (**Table 3**).^15,24,29,38-50^ Assays fall into three mechanistic categories – competitive binding assays, pseudotyped neutralization assays, and live virus neutralization assays. The competitive binding assay measures the ability of antibodies to block interactions between the SARS-CoV-2 receptor binding domain and human ACE2 receptor. Virus pseudotype neutralization assays, mainly HIV- and VSV-based, use full length spike incorporated in the viral particle to measure the capability of neutralizing antibodies to block viral entry into the target cells. SARS-CoV-2 live virus plaque or focus reduction neutralization assays measure the ability of neutralizing antibodies to block the spreading infection of authentic SARS-COV-2 in cell culture. Diagnostic sensitivity and specificity for neutralization methods developed within SeroNet range from 93 – 100 % and 97 – 100 %, respectively. Lastly, 9 institutions report use of 12 commercial serology methods (**Table 4**). Commercial methods detect IgG, IgM, and/or total Ig to spike, RBD, and/or nucleocapsid antigens in serum or plasma. Of the commercial methods in use, 10 are FDA EUA approved, 1 is pending FDA EUA, and 1 is RUO.

**Table 2:**
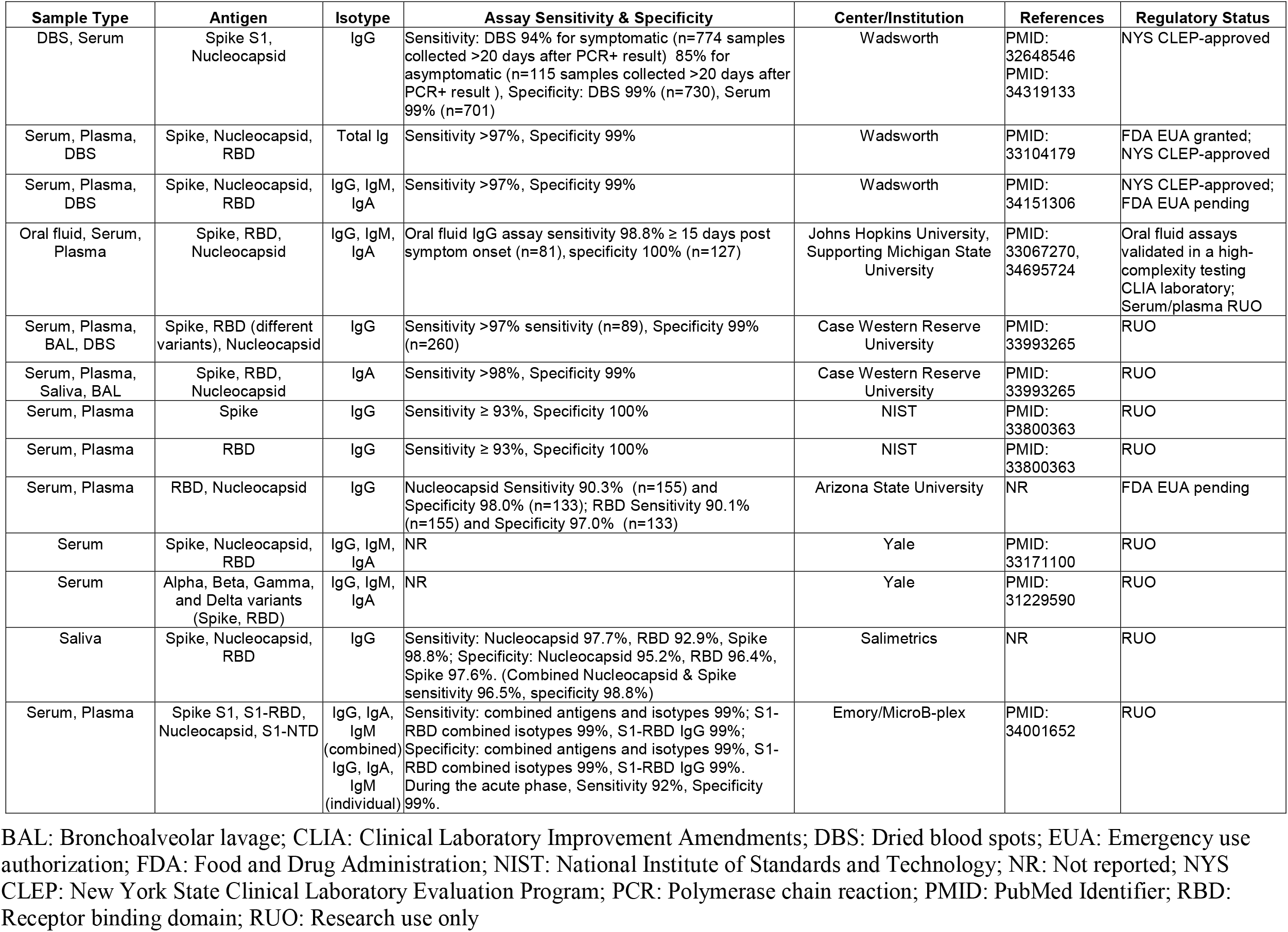
Laboratory-developed multiplex assays

**Table 3:**
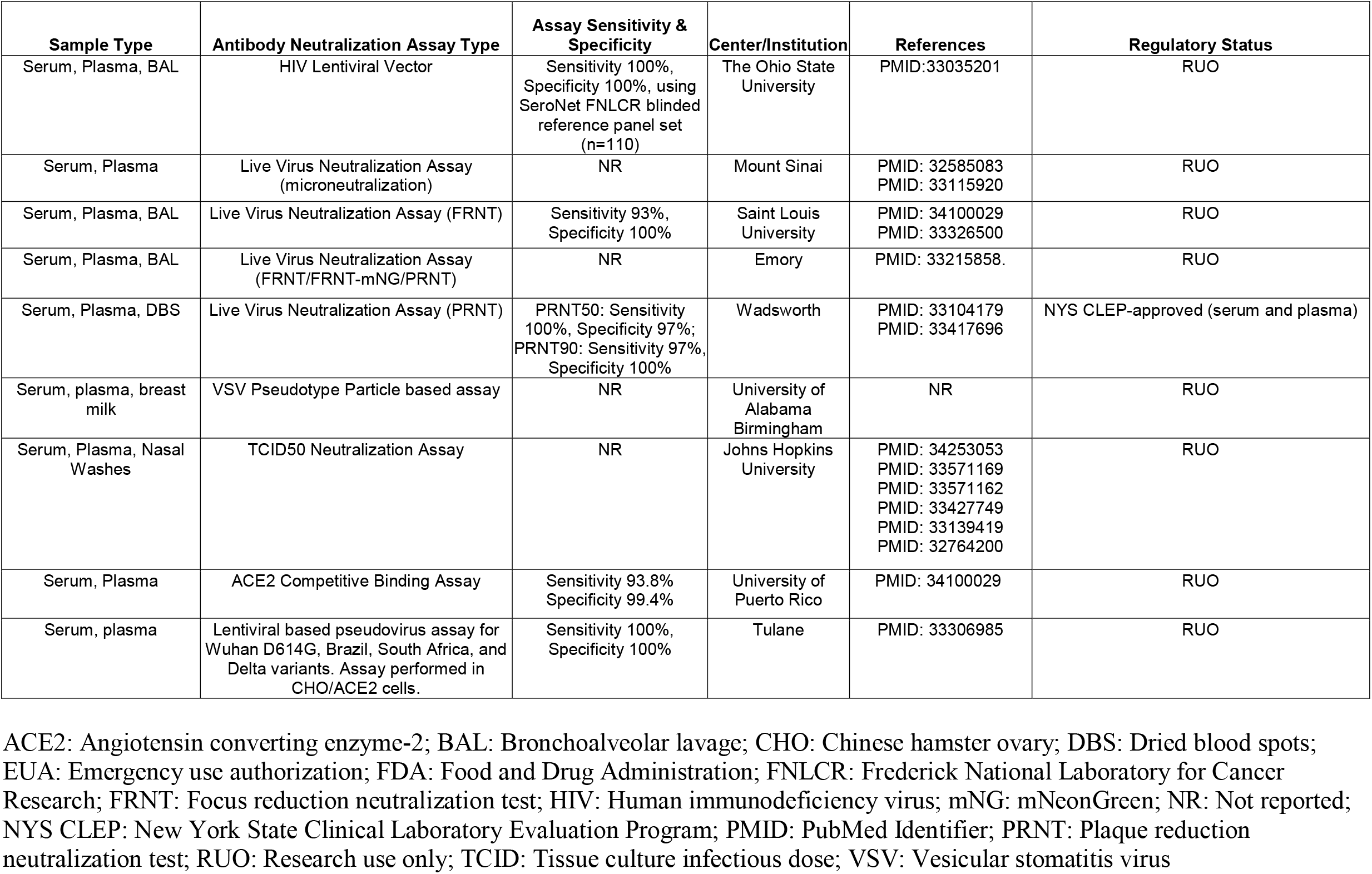
Neutralization assays

**Table 4:** Commercial assays

| **Instrument/Assay** | **Antigen** | **Isotype** | **Center/Institution** | **Regulatory Status** |
| --- | --- | --- | --- | --- |
| Abbott Alinity | Spike | IgM | Mount Sinai | FDA EUA granted |
| Abbott Architect | Spike, Nucleocapsid | IgG | Cedars-Sinai* | FDA EUA granted |
| Beckman Coulter Access | Spike | IgG | Arizona State University | FDA EUA granted |
| Beckman Coulter Access | Spike | IgM | Arizona State University | FDA EUA granted |
| DiaSorin Liaison | Spike | IgG | Feinstein/Northwell, Kaiser, The Ohio State University | FDA EUA granted |
| DiaSorin Liaison | Spike | IgM | Feinstein/Northwell | FDA EUA granted |
| Kantaro SeroKlir | Spike, RBD | IgG | Mount Sinai | FDA EUA granted |
| Kantaro Quantitative SARS-CoV-2 | Spike, RBD | IgG | Mount Sinai | FDA EUA pending |
| Meso Scale Discovery | Spike, Nucleocapsid | IgG, IgM | University of Alabama - Birmingham, CVVR/BIDMC/Harvard, Johns Hopkins University, Stanford | RUO |
| Roche Elecsys Anti-SARS-CoV-2 | Nucleocapsid | Total Ig | University of Minnesota, Feinstein/Northwell | FDA EUA granted |
| Roche Elecsys Anti-SARS-CoV-2 S | RBD | Total Ig | University of Minnesota, Feinstein/Northwell | FDA EUA granted |
| Siemens Atellica | Spike | Total Ig | Kaiser, The Ohio State University | FDA EUA granted |

\*Samples sent to Abbott Diagnostics for testing
BIDMC: Beth Israel Deaconess Medical Center; CVVR: Center for Virology and Vaccine Research; EUA: Emergency use authorization; FDA: Food and Drug Administration; RBD: Receptor binding domain; RUO: Research use only

### Establishment of SeroNet assay traceability to the U.S. SARS-CoV-2 Serology Standard and First WHO International Standard for anti-SARS-CoV-2 Immunoglobulin

Units for the U.S. SARS-CoV-2 Serology standard were initially established by FNLCR based on measurements performed by eight laboratories (**Table 5**). Subsequently, FNLCR further established traceability of the U.S. SARS-CoV-2 Serology standard to the WHO IS 20/136 by using four FNLCR ligand binding serology assays, with assessment of neutralization tested at NIAID’s Integrated Research Facility (IRF) (**Table 5**). The U.S. SARS-CoV-2 serology standard was made available to the public in December 2020. Thus far, there have been 124 requests for U.S. SARS-CoV-2 standard material, and 19 requests for the reference panel samples.

**Table 5:**
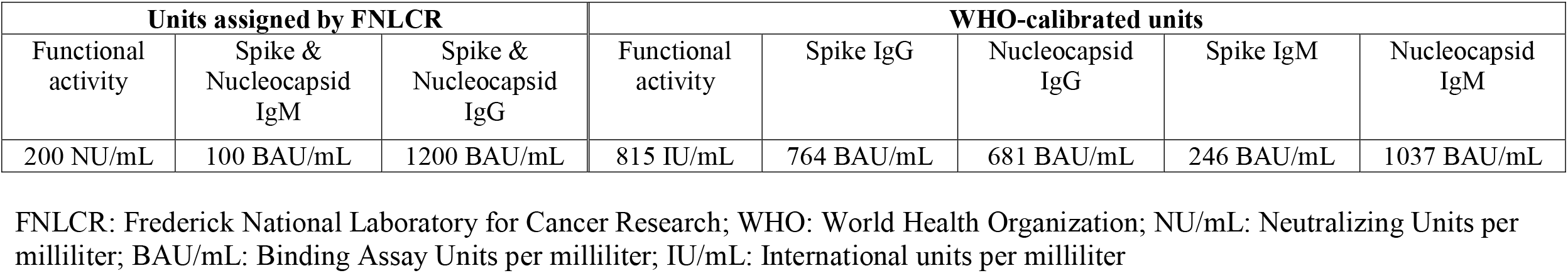
Units assigned to the U.S. SARS-CoV-2 Serology Standard

## Discussion

SeroNet has collectively established a diverse array of methodologies for measurement of SARS-CoV-2 antibodies in a variety of biological fluids. Methods include laboratory-developed ELISAs, multiplex assays, and neutralization assays, most used for research-only purposes, as well as commercial assays available for patient care or research studies. Assays have been developed to test unique sample types, including DBS, saliva/oral fluid, breast milk, nasal washes, and bronchoalveolar lavage fluid. Binding assays identify IgM, IgG, IgA, and/or total antibodies to nucleocapsid, spike, RBD and/or N-terminal domain (NTD) antigens, and neutralization assays rely on three methods to quantify antibodies with functional neutralizing activity. This diversity of assay methods allows for robust investigation of multiple aspects of the serological response to SARS-CoV-2 infection and vaccination, and for cross-comparison of assay performance across platforms and institutions within SeroNet.

With the rapid development of numerous methods for serological assessment, as exemplified by the depth and breadth of assays within SeroNet, it is critical to establish assay harmonization and standardized reporting units to facilitate cross-comparison of results across studies, as well as for streamlined meta-analyses. To this end, FNLCR has provided the U.S. SARS-CoV-2 serology standard reference material, which has traceability to the First WHO International Standard for anti-SARS-CoV-2 Immunoglobulin, to SeroNet sites performing serological assays, to allow establishment of standardized reporting of results in binding antibody units (BAU) per mL traceable to the WHO standard. These efforts may more rapidly facilitate the establishment of a universal cut-off as a correlate of protection, which will be critical to broaden the clinical utility of serological testing for patient care, will allow vaccine trials to transition to an immunogenicity endpoint rather than morbidity or mortality endpoints (immuno-bridging), and will guide decisions regarding optimal scheduling of future vaccine doses to optimize protective efficacy for the general immunocompetent population and susceptible immunocompromised sub-populations.

In summary, SeroNet is well-positioned to rapidly and collaboratively advance our understanding of the immune response to both SARS-CoV-2 infection and vaccination, with ongoing evaluation of serological responses to SARS-CoV-2 variants of concern. The collective effort of institutions involved with SeroNet, to both establish diverse and complementary serological assays, and establish traceability of these diverse assays to the WHO standard, will allow for comprehensive investigation of immune responses and facilitate pooled analyses within the SeroNet consortium. This will enable achievement of the ultimate goal – establishment of a universal correlate of protection cut-off, which will provide a foundation for broader clinical use of serologic testing, as a guide for future decisions on scheduling of COVID-19 vaccine boosters, as well as for general assessment of COVID-19 vaccine immune responses against vaccine viruses and newly evolving variants of concern.

## Data Availability

All data produced in the present study are available upon reasonable request to the authors.

## Funding sources

Funded by NCI Contract No. 75N91019D00024, Task Order No. 75N91021F00001, award numbers 21X089 (J.L., V.M., J.P., J.Q., L.S.), 21X090 (J.M.C., N.C.O.), 21X091 (A.B.K., S.N.T., B.T.), and 21X092 (C.C-C., A.F-B., F.K., D.R.M., V.S., A.W.); and NCI Grants U54CA260591 (K.S.), U01CA260469 (T.L., D.A.G., S.W.G., C.D.H., K.K., N.P.), U54CA260543 (L.P.), U54CA260582 (S-L.L., G.L.), U54CA260492 (S.L.K., S.D.), U54CA260563 (F.E-H.L., M.S.S., N.S.H., J.L.D., J.D.R.), U01CA260541 (J.D.B., A.K.P., T.L.S., E.T.S., C.A.S., P.P., A.M.E.), U01CA260526 (K.W.B., J.C.F., J.L.K.), U01CA261276 (R.A.B., C.F., A.M.M.), U01CA260539 (C.L.K.), U01CA260508 (L.M.S., A.P.D., R.C.G., D.T.H., W.T.L., J.L.Y., A.F.P.), U01CA260462 (S.B., S.P.).

## Potential conflicts of interest

A.B.K. is a consultant for Roche Diagnostics and has received research support from Siemens Healthcare Diagnostics and Kyowa Kirin Pharmaceutical Development. J.D.B., A.K.P., E.T.S., and T.L.S. have received research support from Altimmune. J.D.R. and M.S.S. are co-inventors on a patent filed by Emory University covering the serology assay described in this manuscript. M.S.S. serves on the advisory board for Moderna and Ocugen. F.E.L. is the founder of MicroB-plex, Inc. J.L.D. is the CSO of MicroB-plex, Inc. N.S.H. has been a senior scientist at MicroB-plex, Inc. F.E.L. has research grants from the Gates Foundation and Genentech, is on the SAB of Be Biopharma, Inc., and received royalties from BLI, Inc., as an inventor for the plasma cell survival media. S.B. has research support from Merck and Pfizer, and is a member of the CMV Vaccine Advisory Committees of Merck and Moderna. S.P. has research support from Moderna. D.A.G. is the Chief Scientific and Strategy Advisor of Salimetrics, LLC. S.W.G. is the Chief Scientific Officer of Salimetrics, LLC. Mount Sinai has licensed serological assays to commercial entities and has filed for patent protection for serological assays. The Icahn School of Medicine at Mount Sinai has filed patent applications relating to the COVID-19 serological assay (“Serology Assay”) and NDV-based SARS-CoV-2 vaccines which list F.K. (“Serology Assay”, vaccines), V.S. (“Serology Assay”), A.F-B. (“Serology Assay”), D.R.M. (“Serology Assay”), and C.C-C. (“Serology Assay”) as co-inventors. The foundational “Serology Assay” intellectual property (IP) was licensed by the Icahn School of Medicine at Mount Sinai to commercial entities including Kantaro Biosciences, a company in which Mount Sinai has a financial interest. All remaining authors report no relevant conflicts of interest.

## Notes

### Funding Statement

This study was funded by NCI Contract No. 75N91019D00024, Task Order No. 75N91021F00001, award numbers 21X089 (J.L., V.M., J.P., J.Q., L.S.), 21X090 (J.M.C., N.C.O.), 21X091 (A.B.K., S.N.T., B.T.), and 21X092 (C.C-C., A.F-B., F.K., D.R.M., V.S., A.W.); and NCI Grants U54CA260591 (K.S.), U01CA260469 (T.L., D.A.G., S.W.G., C.D.H., K.K., N.P.), U54CA260543 (L.P.), U54CA260582 (S-L.L., G.L.), U54CA260492 (S.L.K., S.D.), U54CA260563 (F.E-H.L., M.S.S., N.S.H., J.L.D., J.D.R.), U01CA260541 (J.D.B., A.K.P., T.L.S., E.T.S., C.A.S., P.P., A.M.E.), U01CA260526 (K.W.B., J.C.F., J.L.K.), U01CA261276 (R.A.B., C.F., A.M.M.), U01CA260539 (C.L.K.), U01CA260508 (L.M.S., A.P.D., R.C.G., D.T.H., W.T.L., J.L.Y., A.F.P.), U01CA260462 (S.B., S.P.).

### Summary of Updates

Co-authors inadvertently left off the initial submission were added, along with their funding sources and disclosures. Initials of authors added next to funding sources for clarity.

